# Effect of a behavior change intervention on complementary food contamination in rural Bangladesh: a cluster-randomized controlled trial

**DOI:** 10.1101/2024.07.22.24310758

**Authors:** Tarique Md. Nurul Huda, Anna A. Müller-Hauser, Shafinaz Sobhan, Md. Shaheen Hossain, Jesmin Sultana, Mahbubur Rahman, Mohammad Aminul Islam, Om Prasad Gautam, Amanda S. Wendt, Jillian L. Waid, Sabine Gabrysch

**Affiliations:** Department of Public Health, College of Applied Medical Sciences, Qassim University, Buraydah 51452, P.O. Box 6666, Saudi Arabia; Environmental Health and WASH, Health System and Population Studies Division, icddr,b, Dhaka, Bangladesh; Research Department 2, Potsdam Institute for Climate Impact Research (PIK), Member of the Leibniz Association, Potsdam, Germany; Charité – Universitätsmedizin Berlin, corporate member of Freie Universität Berlin and Humboldt-Universität zu Berlin, Institute of Public Health, Berlin, Germany; Global Health and Migration Unit, Department of Women’s and Children’s Health, Uppsala University, Uppsala, Sweden; Paul G. Allen School for Global Health, Washington State University, Pullman, Washington, USA; Food Microbiology Laboratory, Laboratory Sciences and Services Division, icddr,b, Dhaka, Bangladesh; WaterAid UK, London, UK; Heidelberg Institute of Global Health, Heidelberg University, Heidelberg, Germany

**Keywords:** Microbiological contamination, Food hygiene, Complementary feeding, Public health, Bangladesh

## Abstract

Children in low-resource settings often consume microbially contaminated food, posing a risk to their health. We evaluated the impact of a food hygiene intervention on complementary food contamination in Bangladesh. A three-year homestead food production intervention was complemented by an eight-month behavior change module to improve household food hygiene practices and evaluated in a cluster-randomized controlled trial including a dedicated study measuring outcomes along the hygiene pathway to intestinal health. In this analysis, we used multilevel regression to assess the intervention’s impact on microbial food contamination, as well as on food hygiene knowledge (n=518) and reported practices (n=531) among mothers of children aged 6-23 months. Complementary food samples were collected from 342 households with children aged 6-18 months and tested for *Escherichia coli*. Overall, 46% of food samples were contaminated with *Escherichia coli* (42% intervention, 49% control), and there was no evidence that the intervention reduced food contamination (Odds Ratio: 0.7, 95% CI: 0.3-1.2, p=0.18). A higher proportion of intervention mothers could name all key food hygiene practices (22% intervention vs. 0% control), had access to a basic handwashing station near the kitchen (24% vs. 14%, p=0.03), reported washing hands before food preparation and child feeding (21% vs. 8%, p=0.001), washing and storing feeding utensils safely (61% vs. 49%, p=0.02), and preparing food fresh or reheating stored food (88% vs. 79%, p=0.03), compared to control mothers. The intervention thus improved knowledge and reported food hygiene practices among mothers, but this improvement did not result in a substantial reduction of complementary food contamination.

**Trial registration number:** NCT02505711

## Introduction

There has been tremendous progress in child survival, and in the past 25 years, death due to diarrheal diseases has reduced globally ^1^. However, morbidity due to diarrhea has not decreased as fast. Diarrheal diseases continue to be among the leading causes of disability-adjusted life-years (DALYs) in children in 2019 ^2^, and foodborne pathogens often contribute to a significant burden of diarrheal disease ^3, 4^. In addition to causing diarrhea, enteric pathogens transmitted through contaminated food can also cause environmental enteric dysfunction (EED), a subclinical, chronic inflammatory disease of the small intestine which can impact child growth and development ^5, 6^.

Microbiological contamination of food is widespread in low- and middle-income countries (LMICs) ^7, 8, 9, 10, 11^. There is evidence that food contributes more to the pathogen burden for infants than drinking water, as in many studies, the level of enteropathogens found in complementary foods was higher than in drinking water ^12, 13^. Observational studies repeatedly found that poor food hygiene practices, including during preparation, storage, serving and feeding, are associated with microbiological contamination of complementary foods ^7, 14, 15, 16, 17, 18, 19^. Consumption of contaminated complementary foods creates a vicious cycle of diarrhea and malnutrition, threatening the health and development of children ^4, 20^. Despite the high burden of disease associated with contaminated food and the high prevalence of poor food hygiene practices ^7^, there has been a lack of attention to food hygiene in the water, sanitation, and hygiene (WASH) field, as well as in health and nutrition programs.

Several efficacy studies have shown that implementing the Hazard Analysis and Critical Control Points (HACCP) approach can identify risk factors and action points during food preparation, serving, feeding, and storage, which could minimize microbial contamination of infant foods in domestic settings in LMICs ^8, 17, 21, 22, 23^. However, the generalizability of these findings was limited by their short implementation periods (three to four weeks) and the very intensive contact with a small number of study participants (ranging from 10 to 120 households).

Recently, a few intervention studies in South Asia and sub-Saharan Africa attempted to improve caregiver food hygiene practices and thus reduce complementary food contamination by employing social and behavior-change techniques and emotional drivers ^16, 24, 25, 26, 27, 28^. A proof-of-concept study in Nepal used a Behavior-Centered Design ^29^ approach to develop an innovative intervention package targeting multiple food hygiene behaviors and was successful in improving observed food hygiene practices and reducing food contamination ^16, 28, 30^. This behavior-change model was adapted to rural Gambia and replicated in a relatively large community-based program ^24^ and effectively reduced microbial contamination in food and water. A food hygiene intervention in rural Malawi, as part of an integrated WASH project ^25, 27^, improved caregivers’ food hygiene behaviors and reduced diarrhea in children, but did not present results on food contamination. Another food hygiene intervention was conducted in two peri-urban settlements in Kenya, although, to our knowledge, no results have yet been published ^26^. While these studies were of longer duration (three to nine months) and targeted more participants than previous HACCP studies, they still relied on rather intensive interpersonal contact to promote the adoption of food hygiene behaviors. There have been no evaluations of such an intervention at large scale and with less intensity yet. In addition, results on food contamination are only available from two intervention studies so far, in Nepal and The Gambia.

Building upon this background, a food hygiene behavior change module was integrated into a nutrition-sensitive agriculture intervention evaluated in a randomized controlled trial in rural Bangladesh, on a larger scale but with less intensity than previous studies, with the aim to encourage better food hygiene practices among mothers and thus minimize contamination of complementary foods given to their infants and young children. In this article, we assess its effect on reducing contamination in complementary foods, as well as evaluating mothers’ food hygiene knowledge and reported behaviors as intermediate outcomes.

## Methods

### Study design and population

This analysis is part of the “Food Hygiene to reduce Environmental Enteric Dysfunction” (FHEED) study, conducted in the context of the cluster-randomized controlled trial “Food and Agricultural Approaches to Reducing Malnutrition” (FAARM) (ClinicalTrials.gov, ID: NCT02505711). The FAARM trial (2015–2019) evaluated the impact of a homestead food production (HFP) program, implemented by Helen Keller International from mid-2015 to late 2018, on stunting in children under three years of age ^31^. It enrolled 2705 young married women and their children under three years of age from 96 settlements in 13 unions of Baniachong and Nabiganj subdistricts in Habiganj district, Sylhet division in northeastern Bangladesh. A woman was eligible if she reported to be 30 years or younger, was married, had access to at least 40 square meters of land, and was interested in participating in the study, based on a household listing conducted in all villages in the trial area ^31^. Settlements, the unit of randomization, were formed as geographically contiguous areas containing 10 to 65 women, depending on the proximity of residence, with a 400 m buffer between settlements to limit contamination between trial arms.

The homestead food production program promoted year-round homestead gardening, small-scale poultry rearing, and nutrition and hygiene education in woman farmer’s groups. In order to strengthen the food hygiene component of the intervention, we designed a food hygiene behavior change module that was delivered over eight months, from June 2017 to February 2018 ^32^. In doing so, we hoped to decrease food contamination and thus reduce the risk of undernutrition due to intestinal infections. As part of the FHEED study, we assessed the impact of the food hygiene component on microbiological contamination of complementary foods in households with children aged 6-18 months, on food hygiene knowledge and on reported behaviors of mothers with children aged 6-23 months.

### Randomization and masking

Settlements (clusters) were randomized 1:1 using covariate-constrained randomization as described in the study protocol, with 48 clusters allocated to the intervention and 48 to the control group ^31^. The interviewers and the lab staff were not engaged in any of the intervention activities. In addition, they were not informed about the intervention status of the settlements. However, it is possible that the interviewers observed intervention materials, like eye danglers in the kitchen, during data collection. The nature of the study did not allow masking of the implementation team or of participants in intervention settlements. Participants were, however, not made aware that the food sample would be tested for microbiological contamination.

### Development and implementation of food hygiene module

The food hygiene module was adapted from an intervention implemented in Nepal ^16, 30^. Using a Behavior Centered Design approach^29^ the module focused on emotional drivers of hygiene behaviors, such as disgust, nurture, affiliation and pride, and a change in the physical environment, e.g. through placement of visual cues/nudges. The module promoted four food hygiene behaviors: 1) cleanliness of cooking and serving utensils, 2) handwashing with soap and water before food preparation and feeding/eating, 3) safe storage of food and drinking water, and 4) fresh food preparation or thorough reheating of stored food. The behavior change module was implemented among all participant women in FAARM intervention settlements by locally recruited female food hygiene promoters. The food hygiene promoters delivered four attractive and engaging group events and four household visits. A detailed description of the food hygiene module has been published ^32^.

### Sample size

The sample size for this analysis depended on the number of mothers with children in the selected age groups. To assess microbial contamination in complementary food samples, we targeted all mothers with a child aged 6-18 months. Based on the FAARM surveillance system, we identified 402 mothers (intervention: 205; control: 197) eligible to participate in the survey. To assess maternal knowledge and reported practices, we targeted all mothers with a child aged 6-23 months at the time of data collection and identified 685 mothers (intervention: 346; control: 339) eligible to participate in the surveys.

### Data collection

For this analysis, we used data collected as part of the FAARM and FHEED studies: (1) background characteristics collected during the FAARM baseline survey from March to May 2015; (2) complementary food samples and characteristics of these samples, as well as data on environmental spot-checks collected during the cross-sectional FHEED survey from July to September 2018; and (3) data on maternal food hygiene knowledge and reported behaviors collected as part of the routine assessment of the FAARM surveillance system from December 2018 to May 2019.

At baseline, we collected data on household and woman characteristics, such as age, education, household wealth, and religion from all households. We estimated each household’s position within the 2014 Demographic and Health Survey national wealth quintiles according to Equity Tool guidelines using household asset information ^33, 34^. As part of the FHEED cross-sectional survey, we collected samples of children’s complementary foods from all households with a child aged 6 to 18 months and conducted spot-checks to collect information on kitchen and food storage environments of these households (Supplementary Table 1).

Data on reported food hygiene behavior and food hygiene knowledge were collected by adding two questionnaire modules to three consecutive rounds of the FAARM routine assessment (conducted every two months) between December 2018 and May 2019. In the first round, we interviewed eligible mothers on reported food hygiene practices; the reference period was the previous day, questions were not prompted. In the following surveillance round, we asked about food hygiene knowledge. Questions covered general knowledge related to the key food hygiene practices addressed in the food hygiene module. Open-ended, unprompted, questions were used, asking “how” a specific behavior should be practiced and “why” this behavior is essential. In addition, we explored knowledge of five critical time points for handwashing that might directly or indirectly influence food contamination: 1) before food preparation, 2) before eating, 3) before child feeding, 4) after using the toilet, and 5) after touching animals or child feces. Trained interviewers conducted face-to-face interviews using a structured questionnaire. All survey data were collected using the tablet-based Open Data Kit (ODK) application ^35^.

### Collection of complementary food samples

Trained research assistants collected food samples during child feeding, or – in case no feeding event was observed during the household visit – mothers were asked to prepare and serve food as if they would feed their 6-18 months old children. Mothers were asked to place the food in a sterile whirl pack bag with the usual utensils used to feed the child. This was most often by hand. While collecting food samples, the research assistants recorded the temperature of the food using a food thermometer (Manufacturer: SveBake, Model TP500), as well as characteristics of the food, including time of cooking/reheating and duration of food storage. Immediately after collection, food samples were stored in an insulated bag and transported to the food microbiology laboratory of icddr,b within 12 hours of collection, maintaining less than 10 °C temperature at all times. Food contamination was assessed by counting colony-forming units of *E. coli*, a WHO-recommended indicator organism for fecal contamination ^36, 37^.

### Enumeration of *E. coli*

For enumeration of *E. coli*, an aliquot of 25 g solid or 25 ml liquid sample was mixed well with 225 ml of 0.1% peptone water. Samples were diluted in a 10-fold serial dilution until the appropriate number of organisms was obtained. For each food sample, two plates of tryptone bile agar with x-glucuronide (TBX) medium (Oxoid, Basingstoke, U.K.) were inoculated with each dilution by pour plating and incubated at 44 °C for 18 to 24 h according to standard methodology ^9, 38^. The appearance of blue-green colonies on the TBX plate was indicative of the presence of *E. coli* and reported as colony-forming units per gram of food (CFU/g) ^39^. A detailed description is available elsewhere ^40^.

### Outcomes

The primary outcome was microbiological contamination of complementary foods, measured as the percentage of complementary food samples with detectable *E. coli* (≥10 CFU/g food).

Secondary outcomes were the level of *E. coli* contamination in complementary foods, defined as absent/low (less than 10 CFU/g food), medium (10-100 CFU/g food), and high (more than 100 CFU/g food, which is the safety threshold for ready-to-eat foods in microbiological food quality guidelines ^38^), and mean log_10_ transformed *E. coli* counts (log_10_ CFU/g food).

We also assessed caregiver knowledge on food hygiene as well as reported caregiver food hygiene and handwashing behaviors. Reported food hygiene practices are presented as the proportion of respondents who mentioned performing the food hygiene and handwashing behaviors correctly in the past 24 hours (for information on composite indicators see Supplementary Table 1).

### Data analysis

For descriptive tables, we calculated proportions for binary and categorical variables, and means and standard deviations for continuous variables. To assess the effect of the intervention on microbiological contamination as well as on food hygiene knowledge, facilities and reported practice, we compared intervention group outcomes to the control group. We used multilevel models, taking account of clustering by using settlement-level random effects. All analyses are intention-to-treat.

As a sensitivity analysis, we ran a mixed-effects logistic regression model adjusting for the slight imbalance in baseline wealth. We also performed a subgroup analysis, looking at the intervention effect in fresh food and stored food separately. All data analyses were performed in Stata version 15 (StataCorp, College Station, TX, USA).

### Ethical Considerations

The FAARM trial protocol was positively reviewed by Heidelberg University in Germany (Ref.: S-121/2014) and the James P. Grant School of Public Health, BRAC University in Bangladesh (Ref.: 37A). The FHEED study protocol was positively reviewed by Heidelberg University (Ref.: S-606/2017) and icddr,b in Bangladesh (Ref.: PR-17126). All participants provided informed written consent by signature or thumbprint.

## Results

### Characteristics of the study population

We present data from 531 mothers with a child aged 6-23 months at the food hygiene assessment (79% of the 685 mothers eligible). On 518 of these 531 mothers, data on food hygiene knowledge were collected. Food samples were collected from 341 households with children aged 6-18 months (85% of the 402 eligible households). Details about reasons for exclusion are provided in Figure 1.

Baseline characteristics of intervention and control households were largely similar (Table 1). Households had on average 7 members at baseline, around three-quarters of households were Muslim and the remainder Hindu. About two-thirds of households had access to an improved latrine and over half had access to a functional handwashing facility. Almost 90% of mothers had some formal education. There were slightly more households belonging to the poorest wealth quintile in control than intervention settlements (15% vs. 10%).

**Table 1:**
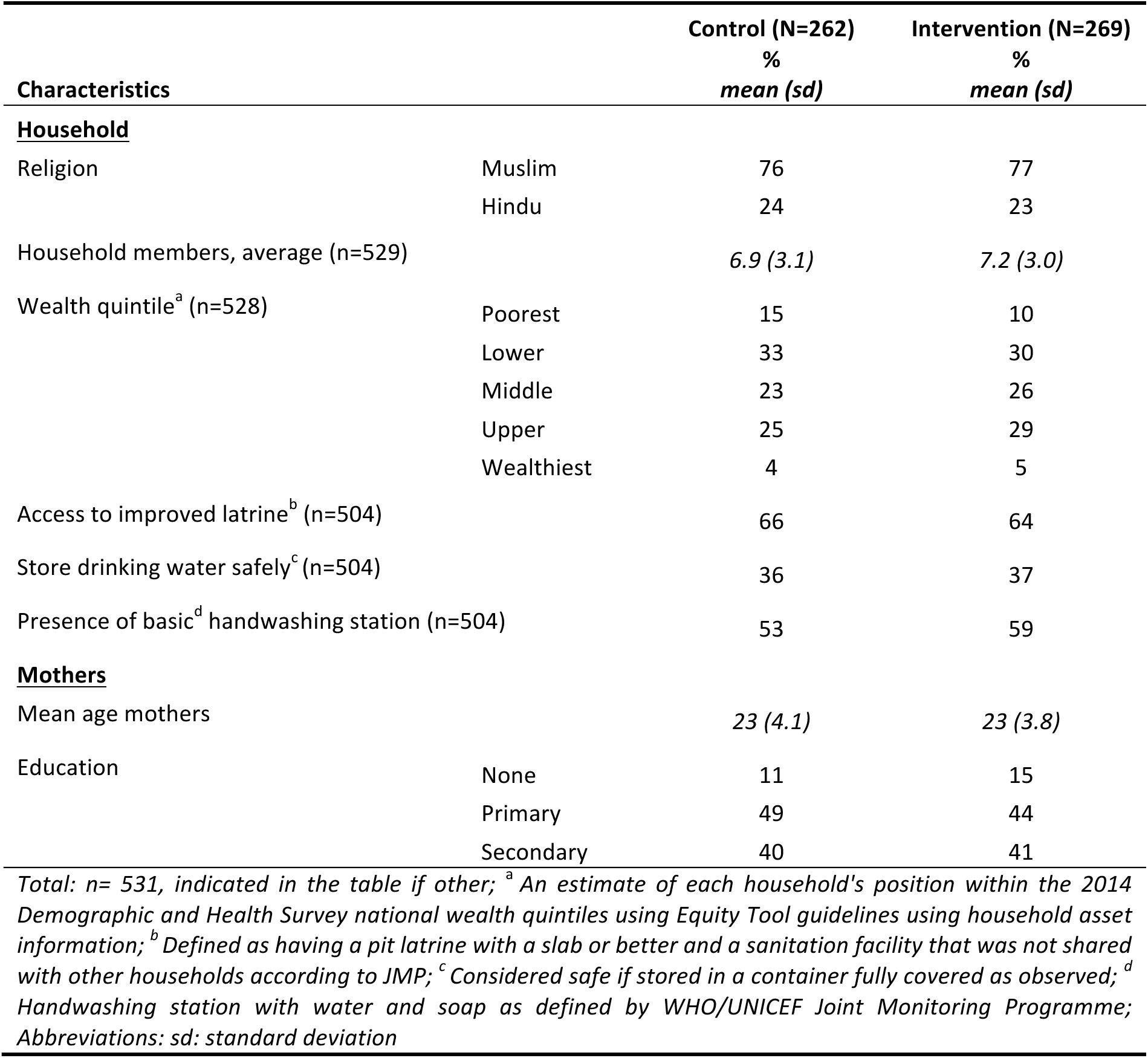
Baseline characteristics of households with mothers of young children among the FAARM study population in rural Sylhet, Bangladesh (2015)

### Impact of the food hygiene intervention on food contamination

Of the 341 food samples tested, 46% were contaminated with *E. coli*. Mean log_10_ *E. coli* counts among all samples were 1.11 CFU/g (SD=1.43) and among *E. coli* positive samples only, the mean was 2.43 CFU/g (SD=1.13). Most foods (88%) had been prepared on the day of sample collection, and around one third of food samples were freshly prepared before child feeding. The most common food was rice (71%), often served with vegetables, pulses, egg, fish, or meat. While there was not much difference in the type of food served, households in the intervention group were more likely to feed freshly prepared food to their children than households in the control group (OR: 1.8, CI: 1.02 – 3.1, Table 2).

**Table 2:** Effect of the food hygiene intervention on food contamination and environmental characteristics.

| Characteristics | Control<br>(N=168) | Intervention<br>(N=173) | OR*/<br>Coef.<sup>#</sup> | CI | p-value |
| --- | --- | --- | --- | --- | --- |
| **<u>Food contamination</u>** |  |  |  |  |  |
| Sample positive for *E. coli* | 49 | 42 | 0.7 | 0.3 - 1.2 | 0.18 |
| *E. coli* contamination: |  |  |  |  |  |
| low (<10 CFU/g) | 51 | 58 |  |  |  |
| medium (10-100 CFU/g) | 18 | 17 |  |  |  |
| high (>100 CFU/g) (vs. medium or low) | 31 | 24 | 0.7 | 0.4 - 1.3 | 0.26 |
| Mean Log<sub>10</sub> *E. coli* counts (CFU/g) among *E. coli* positive samples | 2.48 (1.2) | 2.35 (1.1) | -0.13 | -0.5 - 0.2 | 0.46 |
| **<u>Kitchen and food storage environment</u>** |  |  |  |  |  |
| At least one basic handwashing station near kitchen | 14 | 24 | 1.9 | 1.1 - 3.3 | 0.03 |
| Kitchen area clean | 21 | 17 | 0.7 | 0.3 - 1.4 | 0.32 |
| Food preparation area clean | 19 | 17 | 0.9 | 0.4 - 1.9 | 0.70 |
| Safe storage of food and water | 23 | 27 | 1.2 | 0.7 - 2.0 | 0.48 |
| Mean temperature (°C) of food storage area | 31.0 (1.8) | 31.0 (1.9) | 0.05 | -0.5 - 0.6 | 0.85 |
| Mean humidity of food storage area (in %) | 83.6 (5.7) | 83.7 (5.6) | 0.51 | -1.3 - 2.3 | 0.57 |
| **<u>Food characteristics</u>** |  |  |  |  |  |
| Food prepared fresh | 30 | 42 | 1.8 | 1.02 - 3.1 | 0.04 |
| Type of food: rice vs. other | 73 | 69 | 0.8 | 0.5 - 1.4 | 0.49 |
| Food cooked on day of sampling | 86 | 90 | 1.5 | 0.7 - 3.3 | 0.29 |
| *Total N=341; complementary food has been sampled from children age 6-18 months; \* from mixed-effects logistic regression models with settlement-level random effects. <sup>#</sup> from mixed-effects linear regression models with settlement-level random effects; Abbreviations: sd: standard deviation; OR: odds ratio; Coef.: regression coefficient; CI: 95% confidence interval; CFU: Colony Forming Unit* |  |  |  |  |  |

Slightly fewer food samples from intervention households were contaminated with *E. coli* compared to samples from control households (42% vs. 49%; OR: 0.7, CI: 0.3 - 1.2); however, the evidence that there is an actual underlying difference is rather weak as the observed difference could easily be by chance (p= 0.18, Table 2). Additionally, there were slightly fewer samples with a high grade of *E. coli* contamination in intervention compared to control households (greater than >100 CFU/g: intervention 24% vs. control 31%); yet again, this could be due to chance (p=0.26, Table 2).

After adjusting for wealth in sensitivity analysis, the small difference in food contamination between intervention and control households decreased further (Supplementary table 2). In separate analyses of fresh and stored food samples, there was somewhat less contamination of fresh foods in intervention compared to control households (15% vs. 28%; OR: 0.4, CI: 0.2-1.2), while there was no difference at all in the contamination of stored foods (59% in both, OR: 1.0, CI: 0.5-2.0; Supplementary table 2).

The proportion of households with a clean kitchen or food preparation area was about the same in the intervention and control group and at a low level, at around a fifth. Also, the storage conditions for food and water along with the humidity and temperature of the storage area did not differ. However, intervention households were more likely to have a functional handwashing station in or near their kitchen compared to controls, though still less than a quarter (24% vs. 14%, OR: 1.9, CI: 1.1 - 3.3, Table 2).

### Impact of the food hygiene intervention on mothers’ food hygiene knowledge

Mothers from the intervention group had an overall better knowledge about food hygiene and handwashing practices than mothers from the control group. For example, 19% of intervention mothers could recall five critical time points for handwashing compared to only 5% of control mothers. The intervention especially increased knowledge about handwashing in the context of food preparation (51% vs. 24%), eating (76% vs. 60%) and child feeding (66% vs. 36%) while knowledge on handwashing after defecation was already above 90% and did not increase further (Table 3). Intervention mothers were also more likely to know about other key food hygiene behaviors, namely, to use clean utensils for food preparation (81% vs. 69%), to store food and water safe (48% vs. 23%), and to prepare food fresh or reheat stored foods thoroughly before serving (29% vs. 6%; Table 3). Twenty-two percent of intervention mothers could recall all key food hygiene behaviors, while none of the control group were able to do so.

**Table 3:** Intervention effect on food hygiene knowledge.

| Characteristics | Control<br>% | Intervention<br>% | OR<sup>*</sup> | CI | p-value |
| --- | --- | --- | --- | --- | --- |
| **Knowledge: handwashing at critical time points<sup>a</sup>** |  |  |  |  |  |
| Before food preparation | 24 | 51 | 3.5 | 2.2 - 5.6 | < 0.001 |
| Before eating | 60 | 76 | 2.3 | 1.3 - 4.1 | 0.003 |
| Before child feeding | 36 | 66 | 5.5 | 2.6 - 11.2 | < 0.001 |
| After defecation | 92 | 93 | 1.2 | 0.5 - 2.6 | 0.71 |
| After handling animal/child feces | 49 | 60 | 1.6 | 0.9 - 2.6 | 0.08 |
| All critical times for handwashing mentioned | 5 | 19 | 3.9 | 2.4 - 20.6 | < 0.001 |
| **Knowledge: key food hygiene practices<sup>b</sup>** |  |  |  |  |  |
| Wash hands with soap | 81 | 93 | 4.1 | 1.7 - 10.2 | 0.002 |
| Wash utensils with soap or store at clean place | 69 | 81 | 3.1 | 1.1 - 8.2 | 0.03 |
| Safe storage of food and water | 23 | 48 | 8.0 | 2.9 - 21.8 | < 0.001 |
| Prepare foods fresh or reheat properly | 6 | 29 | 13.0 | 4.3 - 39.4 | < 0.001 |
| All key food hygiene behaviors mentioned<sup>c</sup> | 0 | 22 | - | - | - |
| **Knowledge: correct response regarding the concept of food hygiene<sup>d</sup>** |  |  |  |  |  |
| *Able to describe correct practice of* |  |  |  |  |  |
| Safe food storage | 54 | 65 | 8.1 | 1.4 - 45.3 | 0.02 |
| Reheating leftover foods | 73 | 86 | 2.7 | 1.1 - 6.4 | 0.03 |
| Food hygiene before feeding | 0.9 | 10 | 12.4 | 3.1 - 50.0 | < 0.001 |
| *Able to describe* |  |  |  |  |  |
| Diseases for unsafe food hygiene | 93 | 98 | 5.1 | 0.9 - 29.5 | 0.07 |
| Importance of safe food storage<sup>e</sup> | 98 | 100 | - | - | - |
| Importance of proper reheating | 54 | 70 | 3.8 | 1.2 - 11.8 | 0.01 |
| Importance of cleaning hands and utensils | 90 | 96 | 5.8 | 1.1 - 29.6 | 0.03 |
| *Total N = 518; knowledge has been assessed in households with children 6-23 months of age; knowledge was classified as present if one correct answer – from several possible – was given; <sup>\*</sup> from mixed-effects logistic regression model with settlement-level random effects; <sup>a</sup> Proportion of respondents who could recall (unprompted) critical times for handwashing; <sup>b</sup> Proportion of respondents who could recall (unprompted) key food hygiene behaviors; <sup>c</sup> Due to the limited number of observations, a regression could not be performed; <sup>d</sup> Proportion of respondents who could answer questions related to important concepts of food hygiene correctly; <sup>e</sup> No effect size produced due to insufficient observations; Abbreviations: OR: odds ratio, CI: 95% confidence interval* |  |  |  |  |  |

The intervention also increased mothers’ ability to describe how to correctly practice key food hygiene behaviors (Table 3), namely safe storage (65% vs. 54%) and reheating leftovers (86% vs. 73%), as well as proper hygiene before child feeding – which was at a low level overall (10% vs. <1%). We also assessed mothers’ knowledge on the importance of certain food hygiene behaviors and found it was already above 90% for disease risk, food storage and cleaning of hands/utensils, with marginal improvements. The intervention increased mothers’ knowledge of the importance of proper reheating of stored foods (70% vs. 54%).

### Impact of the food hygiene intervention on mothers’ reported behaviors

Mothers’ reported behaviors were also improved (Table 4), with mothers from the intervention group more likely than controls to report washing hands before food preparation (45% vs. 19%), eating (58% vs. 28%), and child feeding (37% vs. 28%). The intervention also increased reported handwashing after defecation (68% vs. 48%), while there was not much improvement in reported handwashing after cleaning their children after defecation or after disposal of feces, mentioned by less than a third of mothers (Table 4). Moreover, mothers in the intervention group were more likely to report using clean feeding utensils (61% vs. 49%) and preparing foods fresh or reheating stored foods thoroughly (88% vs. 79%), while there was no difference in reported safe storage practices of food (15% vs. 12%) and water (70% vs. 69%; Table 4).

**Table 4:** Intervention effect on reported food hygiene behaviors, early 2019.

| Characteristics | Control<br>% | Intervention<br>% | OR<sup>*</sup> | CI | p-value |
| --- | --- | --- | --- | --- | --- |
| **<u>Behavior: handwashing at critical time points</u>** |  |  |  |  |  |
| Before food preparation | 19 | 45 | 4.0 | 2.3-7.0 | <0.001 |
| Before eating | 28 | 58 | 4.7 | 2.6-8.2 | <0.001 |
| Before child feeding | 18 | 37 | 3.2 | 1.8-5.7 | <0.001 |
| After defecation | 48 | 68 | 2.7 | 1.5-4.8 | 0.001 |
| After cleaning the child's bottom | 18 | 25 | 1.7 | 0.8-3.3 | 0.142 |
| After disposing child's feces | 28 | 33 | 1.5 | 0.7-3.2 | 0.310 |
| **<u>Behavior: key food hygiene practices</u>** |  |  |  |  |  |
| Hands washed before food prep. and feeding | 8 | 21 | 3.5 | 1.7-7.2 | 0.001 |
| Feeding utensils cleaned | 49 | 61 | 1.8 | 1.1-2.9 | 0.021 |
| Food stored safely | 12 | 15 | 1.1 | 0.4-3.4 | 0.832 |
| Water stored safe | 69 | 70 | 1.2 | 0.6-2.7 | 0.573 |
| Food prepared fresh or reheated thoroughly | 79 | 88 | 2.3 | 1.1-4.8 | 0.033 |
| *Total: n=531; reported behavior has been assessed in households with children 6-23 months of age; \* from mixed-effects logistic regression models with settlement-level random effects; Abbreviations: OR: odds ratio, CI: 95% confidence interval* |  |  |  |  |  |

## Discussion

Our trial results of a large-scale, low-intensity behavior change intervention using emotional drivers in two rural sub-districts in Sylhet division, Bangladesh, suggest that the intervention effectively improved knowledge and reported food hygiene practices among caregivers. However, the improvement was insufficient to result in a substantial reduction in microbiological contamination of complementary foods.

The food hygiene module had a high coverage, with more than 75% of women attending at least 7 out of 8 food hygiene sessions^32^. Accordingly, intervention mothers in our study showed overall higher knowledge of almost all food hygiene behaviors than controls, however only 20% remembered all critical times for handwashing and 22% all key food hygiene behaviors. Similarly, while most reported behaviors were higher in the intervention group than in the control group, they were still far from universal. Of the two main food hygiene behaviors linked to a successful reduction in food contamination in FAARM households ^40^, handwashing at critical times was particularly uncommon, with only 21% of mothers in intervention households (vs. 8% in control) reporting that they wash their hands with soap before food preparation and feeding. Fresh preparation of food before feeding was more commonly practiced than handwashing – 42% of food samples collected for food contamination analysis in intervention households were prepared fresh, compared to 30% in control – but also still far from universal. Moreover, the intervention did not increase the low levels of safe food storage and cleanliness of the food preparation and storage areas, which were identified as determinants of food contamination in other studies ^7, 19, 22, 41^, but were less effective in lowering food contamination in FAARM households ^40^.

Since people tend to over-report expected behaviors (social desirability bias) ^25, 42^, actual hygiene practice in study households was likely even lower than reported. In fact, during structured observation of food hygiene behaviors among a subsample of FAARM households with children aged 6-18 months, handwashing with soap was observed in only 12% of intervention households (and 2% of controls), and all recommended behaviors in only 10% of child feeding events ^43^. Food preparation is a multi-step process with several critical points that influence the risk of food contamination/re-contamination ^8^. Therefore, single food hygiene practices have limited potential to prevent contamination, and consistent practice of multiple behaviors is likely needed to substantially reduce food contamination ^40^. In fact, our previous analyses suggest that the most successful single practices (reported handwashing at critical times and observed feeding of fresh food) could each reduce food contamination by about one-third if universally practiced, and food contamination could be reduced by two-thirds if all (reported or observed) food hygiene practices were practiced ^40^.

Taken together, we see an attenuation of both intervention effect and absolute levels from relatively high knowledge of food hygiene to moderate reported behaviors and even lower actual behaviors, which could explain the lack of impact on food contamination found in this study, as well as the lack of impact on environmental enteric dysfunction and diarrhea prevalence that we reported in two previous studies ^44, 45^

In contrast to our findings, two food hygiene studies conducted in Nepal and The Gambia successfully changed several behaviors and reduced microbiological contamination of food ^24, 28^. The higher frequency of exposure and intensive promotional activities in the Nepal and The Gambia interventions may have contributed to their success. In Nepal, 15 promoters delivered weekly intervention activities to 120 mothers over three months, while we had monthly intervention activities delivered by 8 promoters to 1275 mothers over eight months ^30, 32^. In The Gambia, the intervention was delivered to 300 mothers within just 25 days, and additionally included four day-long community campaign events and frequent home visits by community volunteers during that period, with an additional follow-up visit conducted after five months. The intensive village-wide activities and interpersonal visits aimed to change behaviors and social norms ^24^. Our intervention was limited to a smaller number of contact points in order to maintain a workable balance with the ongoing activities of the homestead food production program, as more (or more frequent) visits would have been both difficult to implement and too time-consuming for the study participants ^32^. Given the proximity of intervention and control settlements in our study area, we did not conduct a community-or village-wide campaign to prevent any spillover of intervention activities to controls ^31^.

There is also the consideration of food sampling timing when comparing the results of our study with the other two. We collected food samples four months after the intervention had ended, while in Nepal and The Gambia, samples were collected sooner, within 45 days and one month, of the intervention ending ^24, 28^. It is possible that if we had collected samples immediately after the intervention, we might have seen a greater effect on food contamination. Nonetheless, we wanted to measure the longer-term impact of the intervention and thus evaluated after four months. Moreover, we collected only one food sample per household just before a caregiver fed the child, and at varying times over the day. Therefore, storage time, storage conditions, reheating time, and temperature varied widely. In contrast, the study in The Gambia collected two food samples per household and always in the same way, the first sample immediately after food was prepared in the morning and before feeding the child, and the second sample after storing the same food and before feeding it at lunchtime ^24^. Despite higher food contamination levels in the second (stored) sample compared to the first (freshly cooked) sample in The Gambia, intervention households had substantially lower food contamination levels than control households at both sampling times ^24^. In our study, when we analyzed fresh and stored food samples separately in a subgroup analysis, we also found a reduction in food contamination in fresh food samples – though the evidence for a true difference was again rather weak – but no impact at all on contamination of stored food samples. This indicates that even if the intervention succeeded in improving safe food preparation to some extent, this effect was negated by suboptimal food storage and poor reheating practices, especially as reheating of stored food was overall rarely practiced by study households and – even if practiced – not found to be associated with a reduction in food contamination ^40^.

Finally, it is important to note that enabling technology and supportive infrastructure (e.g. handwashing station with soap and water in or near the kitchen, covering utensils and storage cabinets) can be critical in promoting behavior change and thus reduce food contamination ^46, 47^. We encouraged intervention mothers to reorganize their kitchen and food storage areas to facilitate the practice of the promoted food hygiene behaviors. We also promoted tippy taps for handwashing, but this basic technology was not effectively adopted by households. However, we did not provide any infrastructure such as kitchen sinks to support these changes ^32^. The intervention increased access to a functional handwashing station with soap near the kitchen from 14% to 24%. But this may not be sufficient as still over 75% of households in the intervention group did not have access to a handwashing station with soap near the kitchen, which likely contributed to low handwashing rates. In contrast, in a comparable intervention study in Malawi ^48^, more than half of the intervention households had a handwashing facility with soap and water located close to the kitchen, and in the study in Nepal, more than 50% of study households had piped water on their compounds and about 40% of households had tap water inside the house^30^. Therefore, creating a context-specific environment with enabling infrastructure, like locally acceptable and easily maintainable washing stations in the kitchen with running water, a sink, and proper drainage, that supports the adoption of food hygiene practices will be crucial for the success of future interventions.

Our study has strengths and limitations. To our knowledge, this is only the second food hygiene study (after the one in The Gambia) to implement a behavior change intervention with emotional drivers at scale and assess its impact on microbiological contamination of children’s complementary foods in a randomized controlled trial. It was conducted in rural Sylhet, in a setting that is relatively typical for

Bangladeshi rural villages. The findings thus are likely generalizable to regions with similar demographic characteristics within Bangladesh and other low- and middle-income countries. While we did not have baseline data on food hygiene outcomes, which prevented us from looking at changes over time, the cluster-randomized design with 96 clusters and covariate-constrained randomization should ensure good balance of baseline characteristics, including food hygiene knowledge and practices. When using self-reported data to measure behaviors, social desirability bias is clearly an issue to consider ^25, 42^. However, the very low prevalence of several reported behaviors (e.g. handwashing with soap and safe food storage practices) and the comparisons to structured observation data collected in a subgroup ^43^ give us some confidence that this bias was not large (and for certain behaviors, e.g. using clean feeding utensils, actual practice was in fact better than reported). Most importantly, our main outcome, microbial food contamination, is an objective measure. In combination with previous analyses on observed food hygiene behaviors, environmental enteric dysfunction, and diarrhea prevalence ^43, 44, 45^, our data on food hygiene knowledge, reported behavior, household spot-checks, and food contamination provide us with a detailed understanding of the impact pathway and help comprehend why the food hygiene intervention likely failed to have the desired impact.

We conclude that the intervention based on emotional drivers was successfully implemented at scale and able to improve knowledge and reported practice of food hygiene behaviors but did not lead to a substantial decrease in the contamination of complementary foods, and consequently not to improved intestinal health in children. These findings suggest that in settings with a low level of hygiene infrastructure, changing the physical environment (e.g. access to piped water in the household and/or a kitchen sink with proper drainage) may be needed in addition to hygiene promotion to achieve sustained behavior change at a level sufficiently high to reduce microbiological contamination and improve child health. So, to succeed in the future, food hygiene interventions should consider all necessary contextual, psychosocial, and technological factors.

## Data Availability

A deidentified dataset with the individual participant data that underlie the results reported in this article is available to interested researchers who provide a methodologically sound proposal for use of the data. A data access agreement will need to be signed to gain access to the data.

## Funding source and their role

The FAARM trial was primarily funded by the German Federal Ministry of Education and Research (BMBF, grant number: 01ER1201). The FHEED study was financially supported by a project grant from Deutsche Forschungsgemeinschaft (DFG, German Research Foundation; Project number: 413269709). Foundation Fiat Panis further supported FHEED’s research work. Helen Keller International received additional support from the Carrefour social responsibility program and other charitable donations for implementing the homestead food production program. Sabine Gabrysch received funding through a Recruiting Grant from Stiftung Charité. Funding organizations had no role in the trial design, the intervention or its implementation, in training, data collection, analysis, or interpretation of results.

---

**Supplementary Figure 1:**
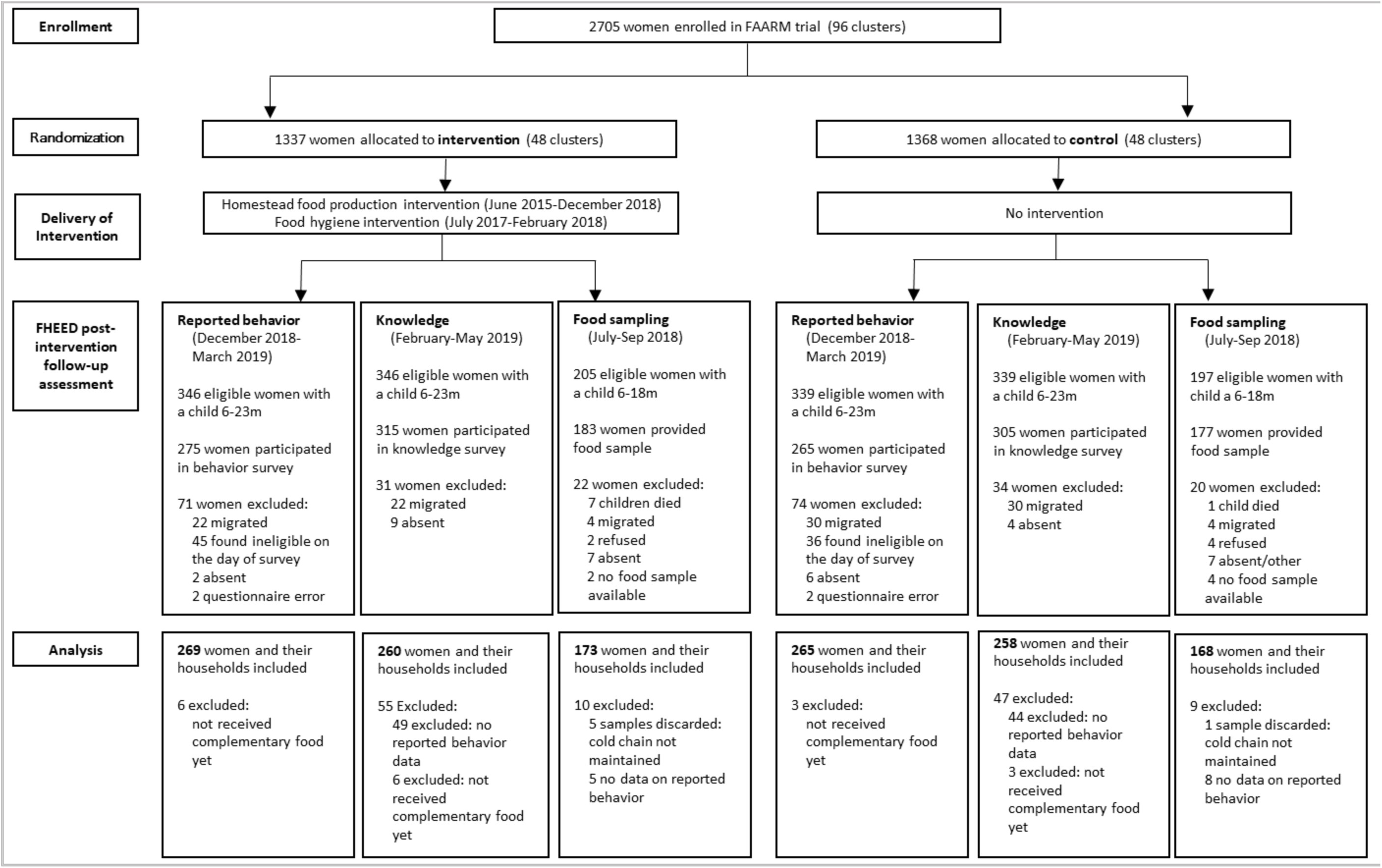
Study Consort flow diagram. FAARM=Food and Agricultural Approaches to Reducing Mal nutrition. FHEED=Food Hygiene to reduce Environmental Enteric Dysfunction.

**Supplementary Table 1:**
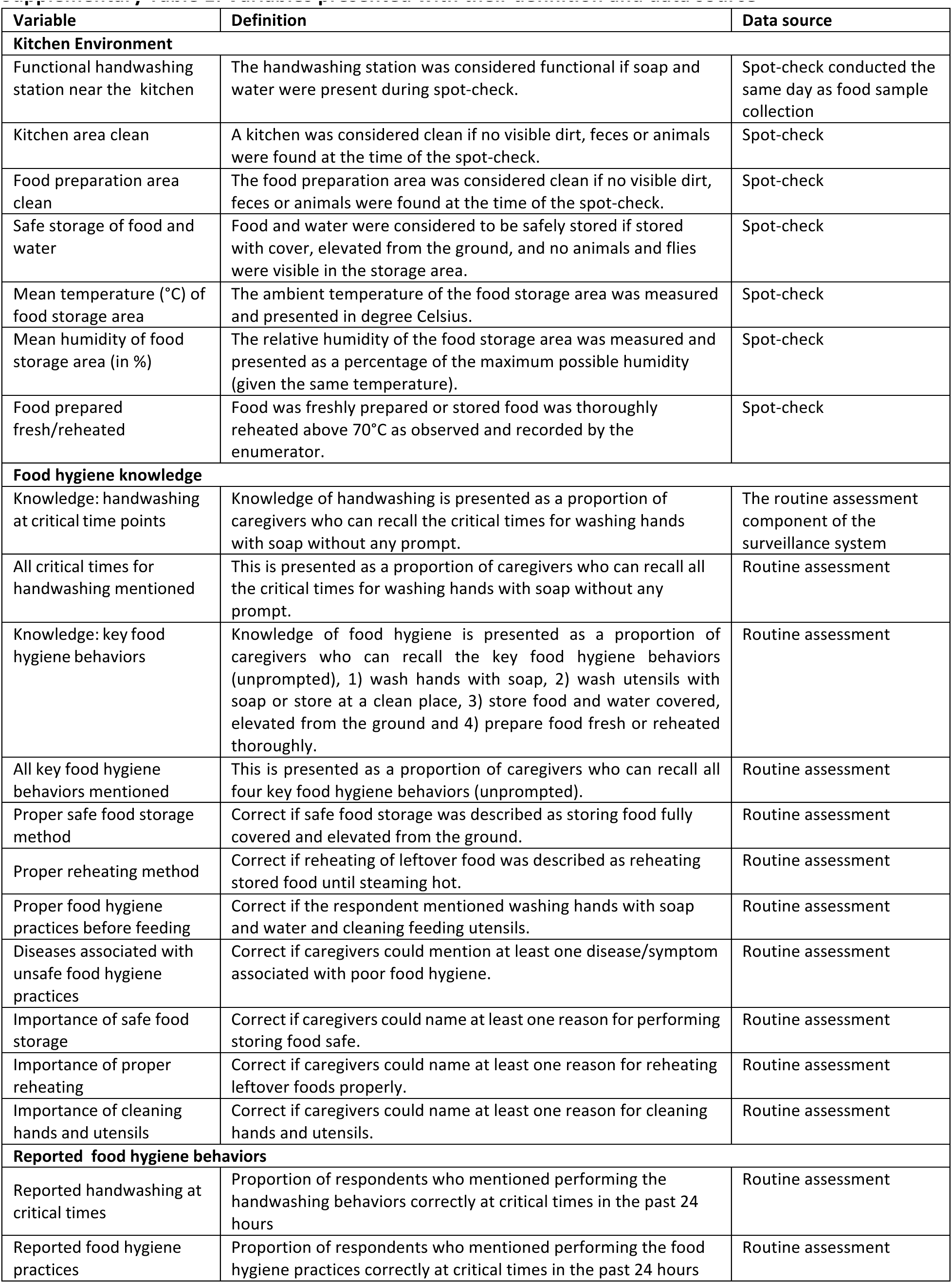
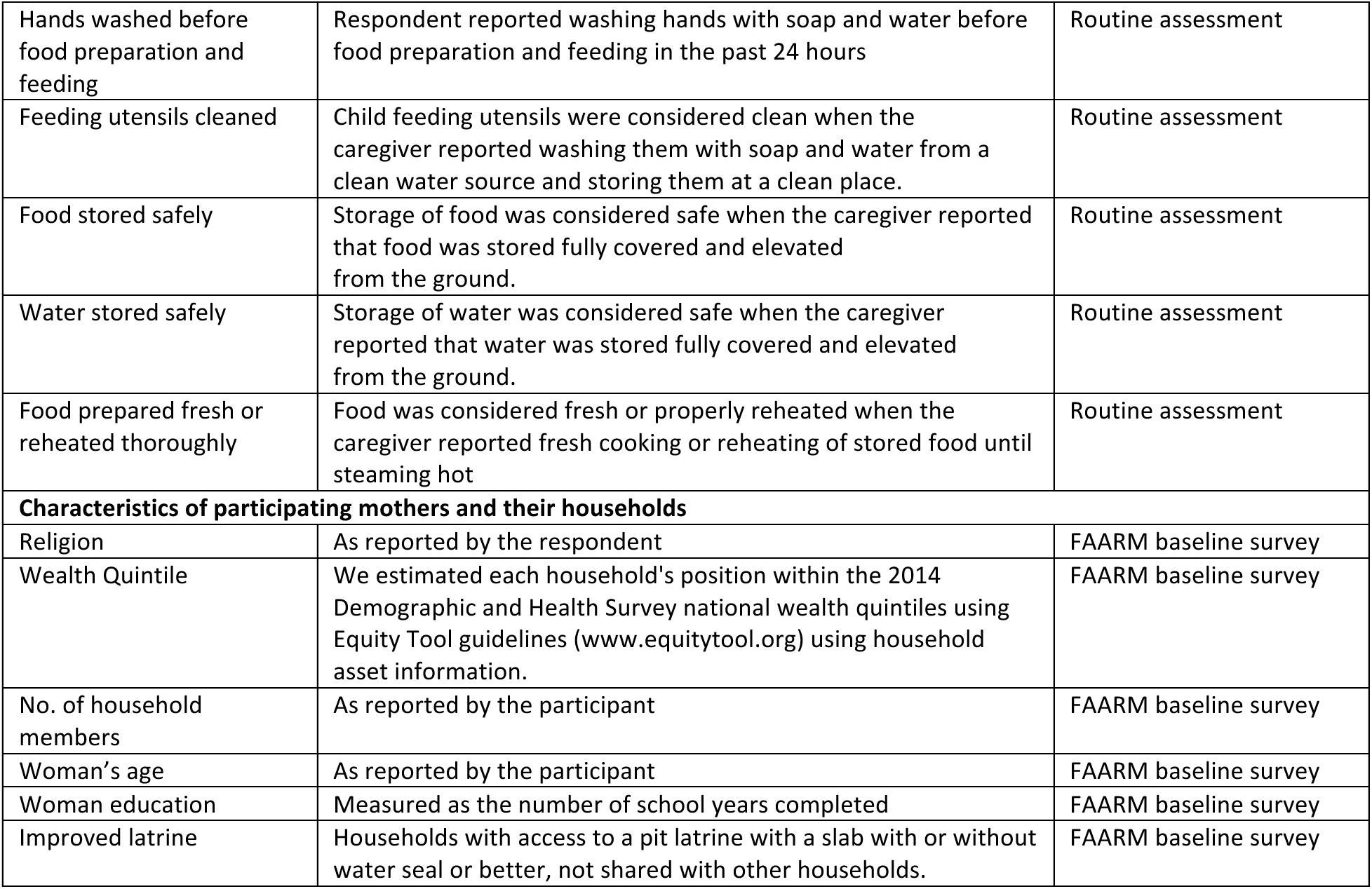
Variables presented with their definition and data source.

**Supplementary Table 2:**
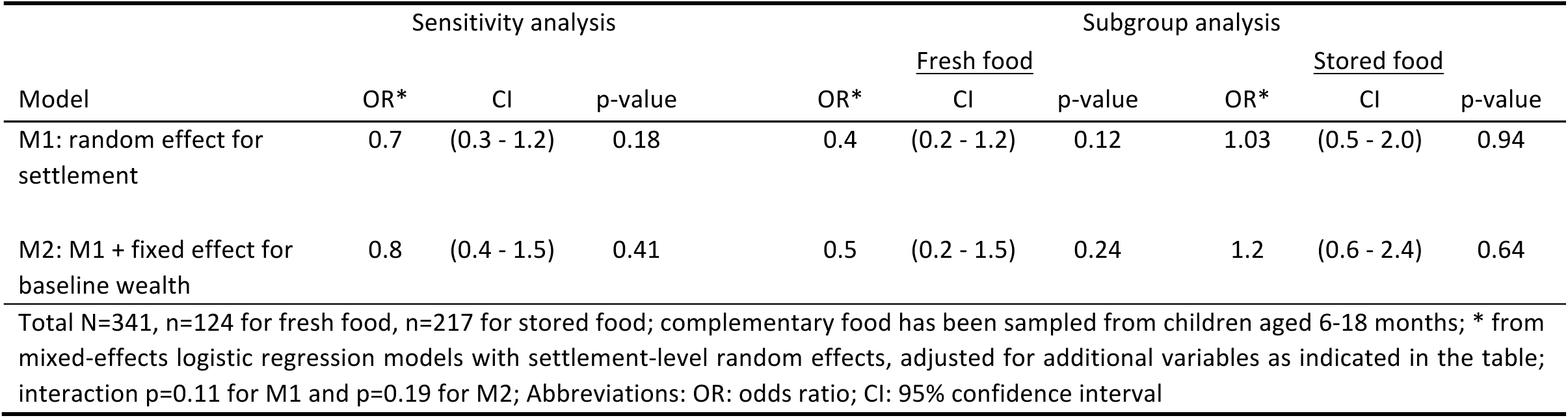
Effect of the food hygiene intervention on food contamination, sensitivity and subgroup analyses.

